# From Unpowered to Actionable: A Bayesian Framework for Evaluating Micro-Cohorts in Graduate Medical Education

**DOI:** 10.1101/2025.11.21.25340770

**Authors:** Carlos Acosta-Batista

## Abstract

**Background:** Graduate Medical Education (GME) programs frequently generate crucial evaluative micro-data from small programmatic cohorts. Traditional frequentist statistics often discard these datasets as “underpowered,” leaving critical curricular gaps undetected. This study demonstrates how Bayesian inference can rescue small-sample educational data to extract robust pedagogical signals.

**Methods:** As a methodological proof of concept, we conducted a Bayesian re-analysis of a historical cross-sectional bioethics evaluation involving a small cohort of Primary Health Care teaching faculty (n=12). A Bayesian Binomial Test with a non-informative Jeffreys prior was employed to calculate the Bayes Factor (BF) and quantify the exact strength of evidence. Evidence was interpreted according to the standard JASP classification guidelines.

**Results:** The Bayesian model extracted high-certainty signals from the historical micro-data. It revealed a stark conceptual dichotomy: Moderate Evidence (BF_+0_ = 8.13) of theoretical mastery in Beneficence, juxtaposed with Extreme Evidence (BF_0+_ = 1320.3) of a complete deficit in the practical application of Non-Maleficence (Prudence).

**Conclusions:** Bayesian inference provides GME leadership with a robust mathematical tool to evaluate small residency and faculty cohorts. By rescuing evaluative micro-data, the model empirically confirms that theoretical teaching does not automatically translate into prudential clinical judgment, justifying targeted curricular interventions even when sample sizes are minimal.

## Introduction

Continuous quality improvement in Graduate Medical Education (GME) relies on Competency-Based Medical Education (CBME), requiring faculty to evaluate whether milestones are attained [1]. However, traditional quantitative and psychometric studies often fail to measure these competencies reliably [2]. Evaluating competency is further challenged by faculty time constraints and decreasing clinical volumes [1].

Programmatic evaluations often involve small, localized cohorts. Traditional frequentist statistics consistently fail in these scenarios due to a lack of statistical power; as Sullivan and Feinn note, sample sizes in GME projects are typically small, often leading researchers to unhelpful non-conclusions simply because their studies lacked the power to detect differences [3]. Methodologically, the p-value relies on imaginary, hypothetical repetitions of data and often overestimates the evidence against the null hypothesis, particularly in small samples [4]. Consequently, valuable evaluative “micro-data” is dismissed.

In contrast, the Bayesian framework conceives of probability as a measure of belief, progressively reducing uncertainty as information accumulates [4,5]. Recent innovations have successfully employed Bayesian frameworks to evaluate rural clinical preceptors [6] and quantify demographic disparities in small programmatic samples [7].

As a methodological proof of concept, this brief report re-analyzes a historical micro-dataset of primary care faculty to demonstrate how Bayesian updating can extract robust, actionable pedagogical signals regarding bioethics competencies from small cohorts.

### Methods: A Worked Example

To illustrate this approach, we utilized a historical baseline dataset comprising a complete primary care teaching faculty cohort (n=12) [8]. The original cross-sectional assessment evaluated five dimensions of bioethical knowledge, with preliminary descriptive findings presented at a local medical education conference [8]. Given the limited sample size, frequentist inference was discarded due to its low statistical power.

To ensure transparency and reproducibility, our analysis and reporting structure adhered to the established guidelines for Bayesian statistical reasoning [9]. We employed a Bayesian Binomial Test utilizing the open-source JASP software (Version 0.19). We tested the hypothesis of adequate knowledge (H_+_: θ > 0.5) against the hypothesis of chance success or null knowledge (H_0_: θ ≤ 0.5). To model initial uncertainty and ensure the objectivity of the re-analysis, a non-informative Jeffreys prior distribution (Beta 0.5, 0.5) was assigned. This methodological choice minimizes researcher bias, ensuring that the resulting Bayes Factor (BF_10_) reflects exclusively the evidence contained in the data [4].

Evidence was interpreted according to JASP’s default classification (BF 3–10: moderate evidence; >100: extreme evidence) [9]. Testing specific, restrictive hypotheses allows for a “severe” test of knowledge, maximizing the discriminatory power of the Bayes Factor even with a minimal sample size [10].

The original data collection for this cohort received formal approval from the Ethics Committee of the “Dr. Carlos J. Finlay” University Polyclinic, with voluntary participation and written informed consent obtained from all physicians prior to data collection. Because the present manuscript is a secondary methodological re-analysis of fully anonymized and de-identified data, it was considered exempt from further Institutional Review Board (IRB) review.

## Results

The Bayesian model successfully extracted high-certainty pedagogical signals from the micro-cohort, demonstrating a marked conceptual dichotomy between the knowledge of traditional principles and their practical clinical application (Table 1).

**Table 1.** Bayesian Proof of Concept: Quantifying the Strength of Evidence in a Small GME Cohort (n=12).

| Questions<br>(Hypothesis $\theta > 0.5$) | Correct<br>Answers<br>(k/n=12) | Proportion<br>(p%) | $BF_{+0}$<br>($H_+$ vs.<br>$H_0$) | $BF_{0+}$<br>($H_0$ vs.<br>$H_+$) | Interpretation of Evidence |
| --- | --- | --- | --- | --- | --- |
| Definition of Bioethics | 2 | 16.7% | 0.069 | 14.49 | **Strong Evidence** in favor of $\theta \leq 0.5$. |
| Bioethics Principles | 4 | 33.3% | 0.110 | 09.09 | **Moderate Evidence** in favor of $\theta \leq 0.5$. |
| Non-Maleficence | 5 | 41.7% | 0.151 | 6.62 | **Moderate Evidence** in favor of $\theta \leq 0.5$. |
| Prudence/Non-Maleficence | 0 | 0.0% | 0.00075 | 1320.34 | **Extreme Evidence** in favor of $\theta \leq 0.5$. |
| Beneficence | 10 | 83.3% | 8.132 | 0.123 | **Moderate Evidence** in favor of $\theta > 0.5$. |

**Table Notes:** **n:** Total number of participants in the cohort. **$\theta$ :** The true underlying proportion of correct answers. **$H_+$ :** The directional alternative hypothesis representing adequate knowledge ( $\theta > 0.5$ ). **$H_0$ :** The null hypothesis representing chance success or lack of knowledge ( $\theta \leq 0.5$ ). **$BF_{+0}$ :** Bayes Factor quantifying the evidence in favor of $H_+$ over $H_0$ . **$BF_{0+}$ :** Bayes Factor quantifying the evidence in favor of $H_0$ over $H_+$ . Evidence interpretation is based on the standard JASP classification guidelines.

For Beneficence, the faculty demonstrated solid theoretical mastery (10/12 correct answers). The model captured this with Moderate Evidence (BF_+0_ = 8.132) in favor of adequate knowledge. In stark contrast, the practical application of Non-Maleficence (Prudence) yielded zero correct answers (0/12). The Bayesian analysis flagged this critical deficit with Extreme Evidence (BF_0+_ = 1320.34) in favor of a lack of knowledge.

Sequential analysis (Figure 1) visually validates this dynamic, demonstrating how Bayesian updating captures real-time evidence accumulation, proving that extreme programmatic certainty can be mathematically established within micro-cohorts.

**Figure 1.**
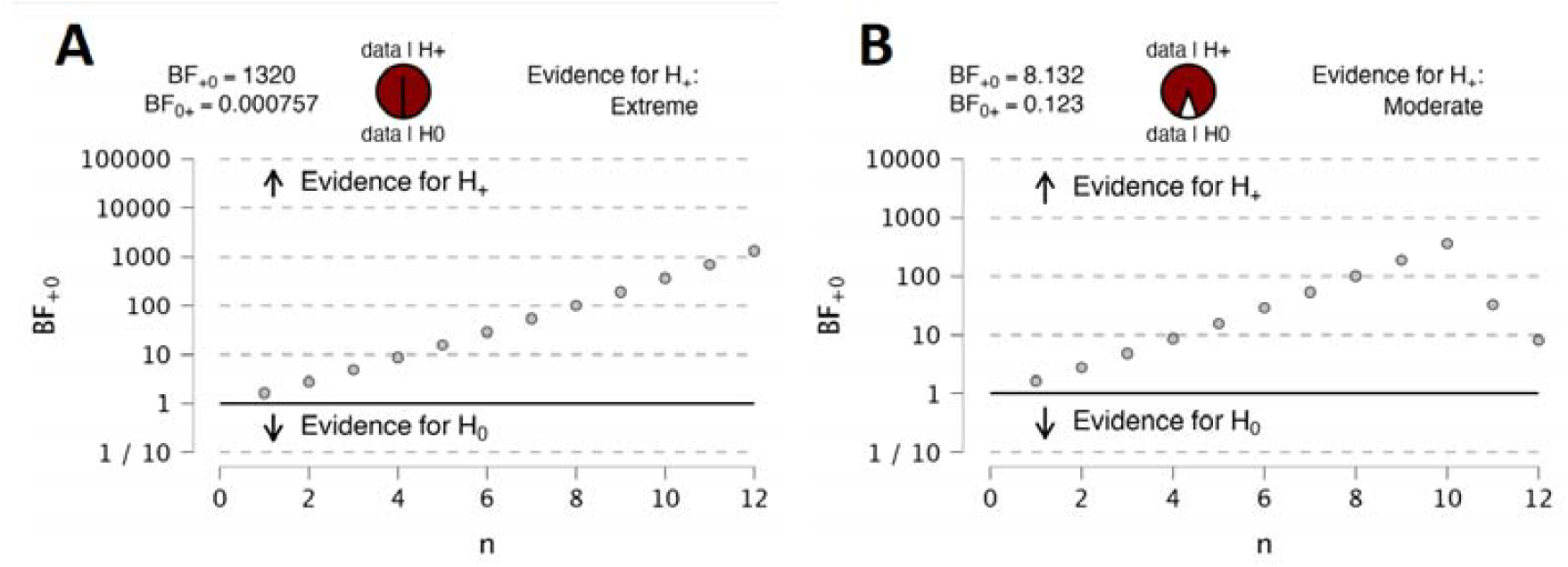
Sequential Analysis of the Bayes Factor in a GME Micro-Cohort (n=12). This diagnostic plot demonstrates how Bayesian updating captures the real-time accumulation of evidence with each added evaluation. **(A) Evidence of a Programmatic Deficit (Prudence):** The trajectory reaches the Extreme evidence level (BF_0+_ = 1320.3) in favor of failure (θ ≤ 0.5), detecting a critical gap in clinical application. **(B) Evidence of Theoretical Mastery (Beneficence):** The plot shows evidence accumulating to achieve the Moderate threshold (BF_+0_ = 8.13) in favor of success (θ > 0.5). As a worked example, these panels prove that actionable pedagogical signals—both positive and negative—can be mathematically established subject-by-subject without relying on large sample sizes.

## Discussion

This proof of concept demonstrates that discarding small-sample GME data as “underpowered” is a missed opportunity [3]. The magnitude of the Bayes Factor (1320.3) allows educators to affirm that they are not dealing with a simple “absence of evidence” due to a small sample, but rather overwhelming “evidence of absence” regarding practical prudential knowledge.

As a methodological note, while software like JASP provides automated heuristic classifications (e.g., Moderate or Extreme) [9], educators evaluating high-stakes micro-data manually can also apply alternative, stringent frameworks, such as the Kass and Raftery (1995) classification scheme, to further calibrate programmatic certainty [11].

The detection of this “Prudence Gap” has profound clinical implications [1]. Bayesian reasoning failures, such as base rate neglect, are a systemic issue in medical education [12]. When primary care clinicians lack a formal framework for prudent clinical judgment (Phronesis) to weigh probabilistic determinants against biomedical utility, it often manifests as subjective hesitation or ethical insecurity in patient care. Fortunately, foundational skills in Bayesian reasoning can be explicitly taught and integrated into faculty development to improve clinical decision-making [13].

This methodological rescue is particularly vital for Family Medicine residency training. Family Medicine programs frequently operate within community-based clinics or rural environments, where resident cohorts and teaching faculties are inherently smaller than those in centralized academic hospitals [3]. Consequently, attempts to evaluate ACGME milestones or assess complex constructs—such as prudent ethical competence in underserved populations—often generate micro-data that traditional frequentist models fail to analyze effectively.

Rescuing this evaluative micro-data allows Program Directors to mathematically validate whether theoretical ethics translates into clinical prudence. This empowers GME leadership to justify targeted curricular interventions without waiting years to accumulate impractically large sample sizes.

The primary limitation of this study is its reliance on a historical, single-center dataset from 2013. Consequently, the specific pedagogical findings regarding bioethics competencies are not broadly generalizable to current primary care cohorts. However, because this study serves as a methodological proof of concept, the primary objective is to demonstrate the utility of the statistical framework rather than to establish universal pedagogical facts. Future research should apply this Bayesian model to contemporary GME datasets across diverse clinical competencies.

## Conclusion

Bayesian inference offers Graduate Medical Education leadership a robust, actionable alternative to frequentist statistics for evaluating small programmatic cohorts. By mathematically validating the certainty of pedagogical signals within micro-data, GME programs can confidently justify targeted curricular interventions without waiting for impractically large sample sizes. This framework is particularly transformative for community-based and rural residency programs, empowering them to rescue valuable evaluative data and drive continuous quality improvement.

## Data Availability

All data produced in the present study are available upon reasonable request to the authors

